# Context Matters! Depression following childbirth or a chronic disease diagnosis shows specific risk factor profiles

**DOI:** 10.1101/2021.10.17.21265109

**Authors:** Bradley S Jermy, Saskia Hagenaars, Jonathan RI Coleman, Evangelos Vassos, Cathryn M Lewis

## Abstract

Progress towards understanding the etiology of major depression (MD) is compromised by its clinical heterogeneity. The variety of contexts underlying the development of a major depressive episode may contribute to such heterogeneity. Here, we aimed to compare risk factor profiles of three subgroups of MD according to episode context.

Using self-report questionnaires and administrative records from the UK Biobank, we characterised three contextual subgroups of MD: postpartum depression (3,581 cases), depression following diagnosis of a chronic disease (409 cases) and a more typical (named heterogeneous) MD phenotype excluding the two prior contexts (34,699 cases). Controls with the same exposure were also defined. We tested each subgroup for association with MD polygenic risk scores (PRS) and other risk factors previously associated with MD (bipolar disorder PRS, neuroticism, reported trauma in childhood and adulthood, socioeconomic status, family history of depression, education).

MD polygenic risk scores were associated with all subgroups, however, postpartum depression cases had higher PRS than heterogeneous MD cases (OR = 1.06, 95% CI: 1.02 – 1.10). Relative to heterogeneous depression, postpartum depression was more weakly associated with adulthood trauma and neuroticism. Relative to heterogeneous depression, depression following diagnosis of a chronic disease did not have higher MD polygenic risk scores but had weaker associations with neuroticism and reported trauma in adulthood and childhood.

The observed differences in risk factor profiles according to the context of a major depressive episode help provide insight into the heterogeneity of depression. Future studies dissecting such heterogeneity could help reveal more refined etiological insights.

## Introduction

Major Depressive Disorder (MDD) is a common psychiatric disorder with a lifetime prevalence between 6.5% - 21% (1). Heterogeneity in the clinical presentation of MDD includes severity and duration of each episode, differential symptomatology, number of episodes experienced, and response to treatment (2). Clinical heterogeneity and a poor rate of treatment response (3,4) argue for subgroups of depression which may benefit from specific treatments and have specific epidemiological and genetic risk factors (5).

Historically, MDD was split into two broad classes, reactive and endogenous depression, which separate patients by a contextual explanation for their depressive episode. Following DSM-III, this separation was removed due to the lack of evidence for differences in outcome between the two diagnoses (6,7). In ‘The Loss of Sadness’, Wakefield and Horwitz argue that the current definition of MDD pathologizes normal sadness by not adjusting the criteria when a state of low mood may be expected, for example following job loss or divorce. Cases in these contexts are less likely to reach the standard of a mental disorder according to a ‘harmful dysfunction’ theory which requires a disorder to result from an impairment to an intrinsic biological or psychological mechanism (8). Indeed, there is evidence that clinicians already consider context when choosing treatment options (9).

Two common contexts behind a major depressive episode are depression in response to a chronic disease and postpartum depression (PPD). The prevalence of MDD in general hospital inpatients is 12% (10) and in PPD is 17% in mothers with no prior history of mental illness (11); both higher than MDD point prevalence in the general population (4.7%) (12).

Many research studies use the term major depression (MD) as a broader definition than MDD, where cases may not be identified using structured clinical interviews and therefore may not attain clinical case status (13). The twin heritability of MD is substantial with estimates ranging between 20% and 50% (14). Genome-wide association studies (GWAS) indicate a highly polygenic architecture for MD, where many common variants of small effect each confer risk (13). Polygenic risk scores (PRS), which capture an individual’s genetic liability to MD, show the genetic relationship between MD and chronic diseases. For example, PRS for MD was not associated with autoimmune disorders despite epidemiological associations (15). In a modestly sized study, PPD, but not MD, was associated with PRS for bipolar disorder (BPD) suggesting a possible biological relationship between BPD and PPD (16).

Being diagnosed with a chronic disease or having a new-born baby both require a sudden adaptation to a new environment. A chronic disease may require learning to live with an increased level of disability and possible facing one’s mortality; the postpartum period requires taking on responsibility for another human life, compounded with sleep deprivation. A period of sadness could be expected in both instances, and a milder version of PPD, ‘baby blues’, occurs in approximately 85% of mothers (17). Wakefield and Horwitz’s harmful dysfunction theory leads to the hypothesis that these subgroups of MDD may have distinct biological and psychological mechanisms and might be less influenced by genetic risk factors.

In this study, we defined MD by context of the depressive episode in UK Biobank (18). We stratify MD cases into three groups: depression diagnosed after diagnosis with a chronic disease, postpartum depression and a typical heterogeneous definition of MD excluding the two prior contexts. To dissect the differential risk factor profiles for these MD subgroups, we characterised their associations with known MD epidemiological risk factors and polygenic risk scores.

## Methods

### Data

The UK Biobank recruited 502,655 participants aged 40-69 in 2006-2010 (18). To define MD cases and subgroups, we combined self-report data - nurse interviews at baseline assessment and the Mental Health Questionnaire (MHQ) completed online by 157,366 participants in 2016-17 (19) - with administrative data from Hospital Episode Statistics (HES) and primary care records. Full details are given in Supplementary Methods.

### Heterogeneous Depression

MD cases were defined using the CIDI-SF within the MHQ and/or a primary care code specific to depression. Cases were allocated to the contextual MD definitions of postpartum depression and depression following diagnosis of a chronic disease, then heterogeneous depression was defined as MD cases not allocated to either context (Figure 1a).

**Figure 1:**
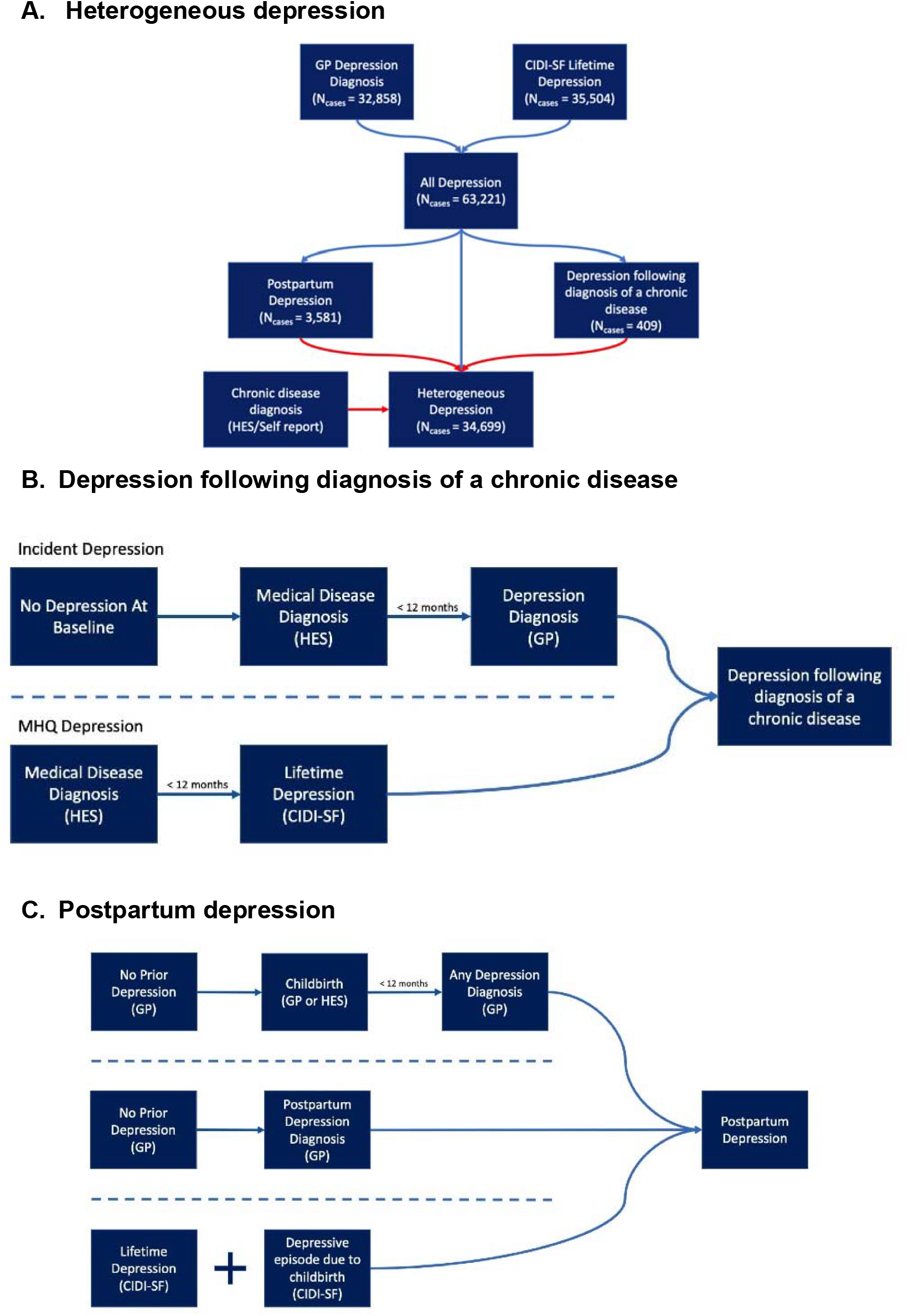
Defining the contextual definitions of major depression. **A.) A flowchart for the three definitions of major depression used in the study**. All diagnoses represent the union of two major depression definitions from self-report and primary care (GP) data. Cases are then allocated to one of three definitions: postpartum depression, depression following diagnosis of a chronic medical disease, heterogeneous depression. Arrows in red represent exclusion criteria. Cases of postpartum depression as well as any cases that report a chronic disease considered in this study are therefore removed from the definition of heterogeneous depression. **B.) A summary schematic of the requirements a participant must meet to be designated case status for depression following diagnosis of a chronic disease. C.) A summary schematic of the necessary requirements a participant must meet to be designated case status for postpartum depression**. In both figures, arrows represent the flow of time and dashed lines separate qualifying criteria. For both depression following diagnosis of a chronic disease and postpartum depression only one set of criteria separated by the dashed lines is required to attain case status. Abbreviations: GP – General Practitioner (refers to primary care data), HES – Hospital Episode Statistics, CIDI-SF – Composite International Diagnostic Interview Short Form (taken from the mental health questionnaire).

### Depression Following Diagnosis of a Chronic Disease

This phenotype is defined as participants experiencing their first major depressive episode (MDE) within a year of receiving a diagnosis of a chronic disease. We selected a subset of diseases associated with increased risk of MD: Stroke (20), Diabetes (21), Autoimmune Disorders (22), Cancers (excluding non-melanoma skin cancers) (23), Motor Neurone Disease (24), Myocardial Infarction (25) and extracted the date of first diagnosis (Supplementary Table 1).

The final phenotype represents the union of cases across two definitions – incident depression and MHQ depression. *Incident depression* cases were participants who do not report MD at baseline and subsequently had their first MD diagnosis according to administrative data (Supplementary Methods). Cases were retained if depression onset was within 12 months of a chronic disease diagnosis (Figure 1b).

*MHQ depression* cases were defined using the CIDI-SF (26) following DSM criteria for an individual’s worst episode of depression. The reported age of their first MDE was matched against the age at onset for each of the chronic diseases. Cases were retained if the two ages were equal or the first MDE was one year later.

### Postpartum Depression

PPD cases were identified from three UKB data sources (Figure 1c).

1. MD cases from the MHQ CIDI-SF who confirmed the worst depressive episode was ‘possibly related to childbirth’ (field 20445).
2. A primary care record for PPD (read codes: 62T1, Eu530) with no earlier records of depression.
3. In primary care records, the earliest date of MD diagnosis was within one year of giving birth.

#### Definition of Controls and Exclusion Criteria

For each depression subgroup, we defined controls having the same exposure as the cases: controls for PPD must have given birth, controls for depression following a chronic disease must have been diagnosed with one of the chronic diseases, and controls for the heterogeneous definition of depression must not have reported any of the chronic diseases. Control groups were screened for multiple definitions of MD. Cases and controls were excluded if they reported a diagnosis of, or medication for, psychosis, bipolar disorder, or substance abuse.

### Risk Factors

Six epidemiological risk factors previously associated with MD were selected (27): education, socio-economic status (SES), neuroticism, family history of severe depression (28), reported childhood trauma and reported adulthood trauma. Education is a four-level categorical variable with qualifications attained at (1) college or university [reference group], (2) age 18 (A-levels’ or equivalent), (3) age 16 (GCSE or equivalent) and (4) no reported qualifications. SES was measured using the regional Townsend Deprivation Index (TDI) and neuroticism was the sum score of a 12-item neuroticism questionnaire. For trauma variables, items from a 16-item trauma questionnaire were assigned to childhood or adulthood. Summary trauma measures were defined as the first principal component of the tetrachoric correlation matrix, separately for childhood and for adulthood reported trauma. These components explained 55.0% and 27.5% of the variance in reported childhood and adulthood trauma respectively (Supplementary Figure 2).

For PRS, a previously defined pipeline for genetic quality control (QC) was applied to retain unrelated participants of European ancestries (18,29). PRSice v2 (30,31) was used to generate PRS for MD and BPD using summary statistics from Wray et al. (13) (N_cases_=116,404, N_controls_=314,990; UK Biobank samples removed) and Stahl et al. (32) (N_cases_=20,352, N_controls_=31,358) respectively. P-value thresholds (P_t_) previously shown to optimise prediction were selected (MD PRS P_t_ < 0.05; BPD PRS P_t_ < 0.01) and a sensitivity analyses was performed including all SNPs (P_t_ <= 1).

### Statistical Analysis

Complete case logistic regression was used to test for association between case-control status under each MD definition and each risk factor. For analysis of epidemiological risk factors, covariates of assessment centre, education, TDI and year of birth were included. Six genetic principal components, genotyping batch and assessment centre were included as covariates for PRS regressions. PRS, neuroticism and TDI were standardised prior to analysis.

To determine if the effect size for a risk factor differed between MD subgroups, odds ratios for PPD and depression following a chronic disease were compared with heterogeneous depression using the Wald test.

A case-case analysis was performed to understand heterogeneity between MD cases, comparing the heterogeneous definition of depression with PPD and with depression following a chronic disease for each risk factor.

Sensitivity analyses were performed by source of MD diagnosis from primary care or self-report (Supplementary Figure 1; Supplementary Table 4), and for PPD using only female participants for the heterogeneous definition.

The Benjamini-Hochberg false discovery rate (FDR) method was used to correct for multiple testing (q < 0.05) (33). Correction was performed separately for the set of case-control, case-case and female-only regressions and Wald tests.

## Results

### Phenotypes

Analysis of UK Biobank identified 3,581 cases for PPD, 409 cases for depression following a chronic disease, and 34,699 cases for heterogeneous depression. Sample characteristics of each group are shown in Table 1, with risk factor missingness and self-reported ethnicity in Supplementary Tables 5-6.

**Table 1:**
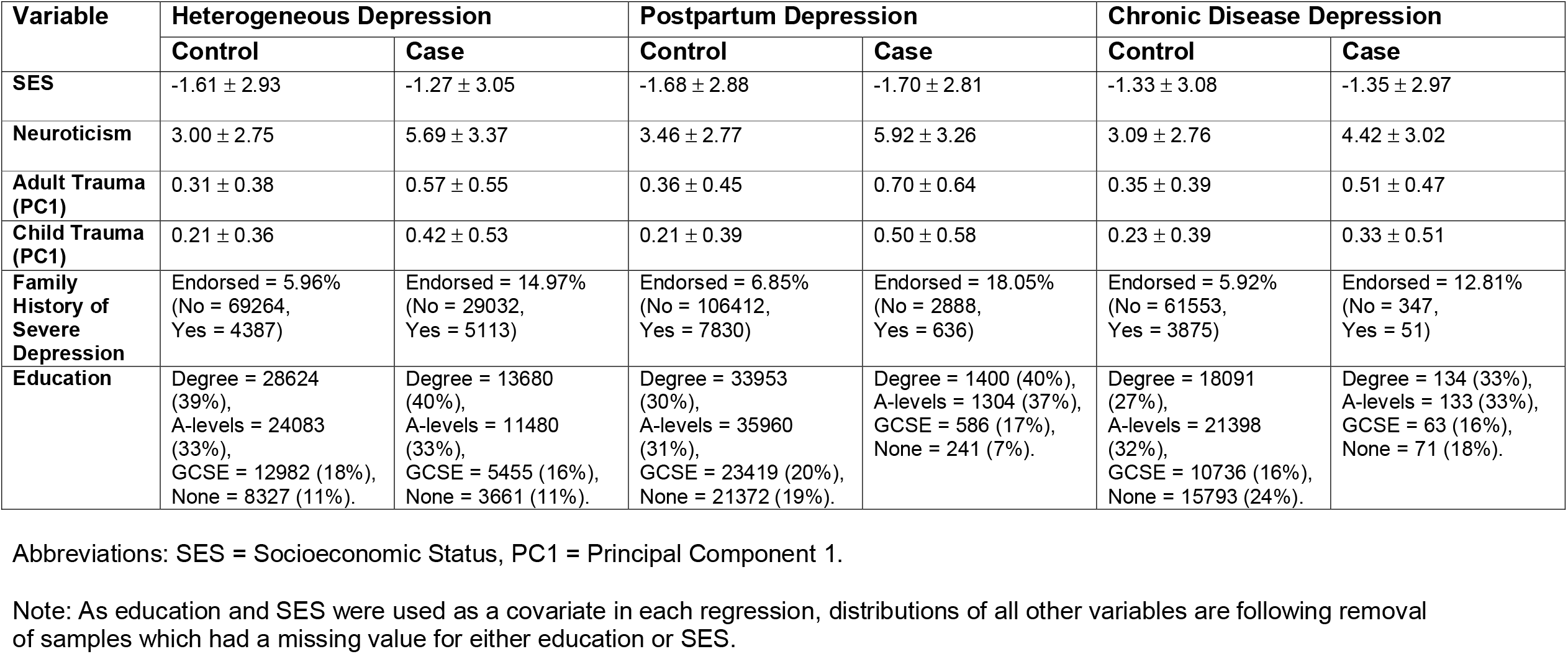
Distribution of values in epidemiological and genetic risk factors across three definitions of MD. Continuous variables have been reported using the mean and standard deviation of the distribution. Binary and categorical variables are reported as percentage endorsed. Values reported represent the raw scores (variables have not been standardised).

### PRS

All PRS were standardised within each group, so odds ratios (OR) represent the change in odds of being a case per one standard deviation increase in PRS. MD PRS was associated with each depression phenotype compared to controls (heterogeneous depression: OR = 1.25, 95%CI = 1.23 – 1.27; depression following a chronic disease: OR = 1.18, 95%CI = 1.05 – 1.31; PPD: OR = 1.29, 95%CI = 1.24 – 1.34; Figure 2a). Relative to heterogeneous depression, PPD and depression following a chronic disease had comparable effect sizes.

**Figure 2:**
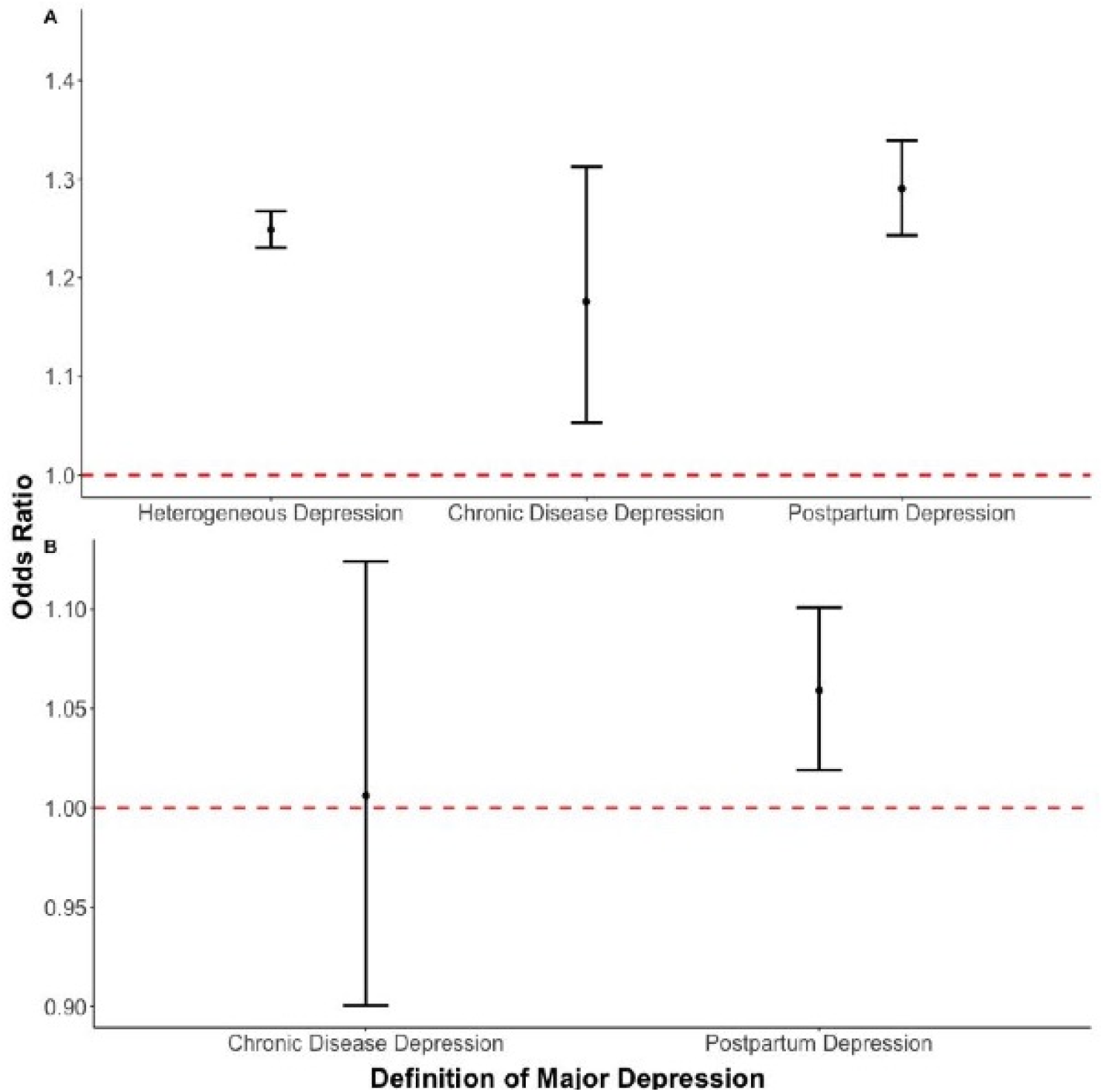
**A) Associations of major depression polygenic risk score with three contextual subgroups of major depression. B) Association of major depression polygenic risk score comparing cases of postpartum and depression contexts of a depressive episode to a heterogeneous definition of depression**. In each test, heterogeneous depression is the reference group. Error bars in both panels represent 95% confidence intervals. The dashed red line represents the point at which the risk factor shows no association with the contextual definition of major depression (odds ratio = 1).

BPD PRS was also associated with each depression definition, with comparable estimates across groups (heterogeneous depression: OR = 1.09, 95%CI = 1.07 – 1.11; depression following a chronic disease: OR = 1.16, 95%CI = 1.04 – 1.30; PPD: OR = 1.09, 95%CI = 1.05 – 1.14; Supplementary Figure 3). Similar effect size estimates were obtained from the sensitivity analysis using PRS with all SNPs (Pt <= 1) (Supplementary Figure 4).

We performed case comparisons to determine if the context of a depressive episode indexed heterogeneity within cases for MD and BPD PRS. The case-only analysis removes variation between our control groups and therefore has a different interpretation from the case-control analyses above. MD PRS was associated with PPD compared to heterogeneous depression (OR = 1.06, 95%CI = 1.02 – 1.10; Figure 2b). Associations with PPD remained significant after restricting heterogeneous depression to females only (Supplementary Table 7). No differences were found between groups for BPD PRS (Supplementary Table 8; Supplementary Figure 5).

### Risk Factors

In general, the risk factors were significantly associated with each MD definition. Depression following a chronic disease was not associated with SES, as measured by TDI, or education. PPD was not associated with SES (Figure 3a-b; Supplementary Table 9).

**Figure 3:**
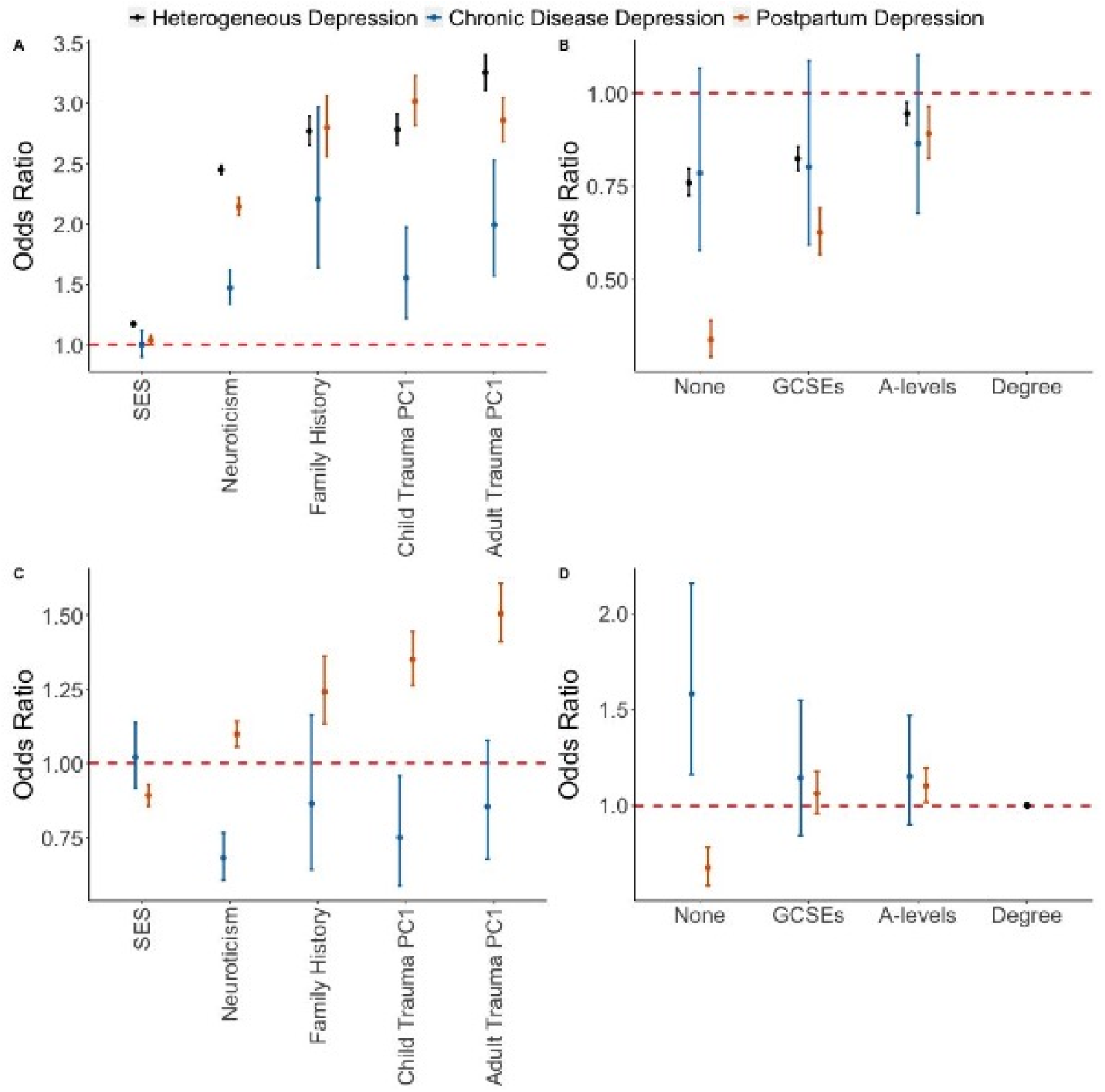
**A) Association of three contextually based subgroups of major depression with epidemiological risk factors. B) Association of three contextually based subgroups of major depression with education**. Having a college or university degree is the reference category. **C) Case-case comparisons for epidemiological risk factors. D) Case-case comparison for educational attainment with all individuals with a university degree as the reference**. In each case-case comparison, cases of heterogeneous depression are the reference group. For all graphs error bars represent 95% confidence intervals. The dashed red line represents the point at which the risk factor shows no association with the contextual definition of major depression (odds ratio = 1). Supplementary figure 6 displays the same associations using log odds to allow for a linear comparison between risk factors and their differences between contextual subgroups.

#### Depression following a chronic disease

Relative to heterogeneous depression, neuroticism, SES and reported childhood and adulthood trauma had weaker associations with depression following a chronic disease (Figure 3a; Supplementary Table 9). Comparing cases of depression following a chronic disease to heterogeneous depression revealed the former case group had lower neuroticism scores, were less likely to report childhood trauma and were more likely to report not having any qualifications compared to a degree (Figure 3c and 3d).

#### Postpartum Depression

The ORs for SES, neuroticism and reported adulthood trauma were lower for PPD relative to heterogeneous depression. Higher education was associated with greater risk of PPD, with a larger dose response shown relative to heterogeneous depression (Figure 3a and 3b). In PPD, the differences in odds ratios for reported adulthood trauma did not survive multiple testing in analysis of the female subset of heterogeneous depression (Supplementary Table 10).

PPD cases showed increased neuroticism scores, family history of severe depression, and reporting of childhood and adulthood trauma compared to cases of heterogeneous depression. PPD cases had higher SES, were more likely to have attained A-levels and less likely to report no qualifications (Figure 3c and 3d). The association with A-levels in PPD was attenuated when the analysis was repeated using female-only heterogeneous depression. While most odds ratios remained consistent in this re-analysis, reports of adulthood trauma reduced from 1.51 to 1.26 (Supplementary Table 7).

## Discussion

We sought to understand the role of context on depression by showing differential associations with genetic and epidemiological risk factors in three MD subgroups - depression following diagnosis of a chronic disease, postpartum depression, and a non-contextual definition – named, heterogeneous depression.

Risk factors of MD PRS, BPD PRS, neuroticism, family history of severe depression and reported trauma in both childhood and adulthood were associated with MD in each subgroup. Other epidemiological risk factors showed a more complex pattern: SES was associated with both PPD and a heterogeneous definition of MD whereas education was associated with heterogeneous MD at all levels and PPD for most levels (Supplementary Discussion). The association across contextual subgroups suggests these risk factors remain important to the etiology of MD, independent of the context of diagnosis. If MD PRS becomes sufficiently predictive to be implemented as a clinical decision tool for depression, the comparable associations across contextual subgroups suggests there will be a minimal discrepancy in utility. However, this requires our findings to be replicated in future, more powerful, MD PRS.

Depression following a chronic disease had weaker associations with SES, neuroticism, and reported adulthood and childhood trauma, compared to heterogeneous depression. Low SES is associated with poorer physical health and mental health (34). Controls with a chronic disease had lower SES than controls for heterogeneous depression (Supplementary Table 11), reducing the association between SES and depression following a diagnosis of a chronic disease.

Trauma and neuroticism may play a weaker role in depression when faced with the immediate stress of being diagnosed with and living with a chronic disease (Supplementary Discussion). This assumes that the chronic disease is the primary reason for the depressive episode and would imply disease related risk factors for this depression subgroup. Indeed, it has been shown that a patient’s level of disability after a stroke (35), physical capabilities among patients with type 2 diabetes (36) and disease course in multiple sclerosis (37) are all associated with MD.

PPD showed a weaker association of reported adulthood trauma compared to a heterogeneous definition of depression. The difference may be driven by the female-only control group. Women are more likely to report interpersonal traumatic experiences, including sexual assault (38) which is an important component of adulthood trauma. PPD controls had higher levels of reported adulthood trauma than heterogeneous depression controls (Supplementary Table 11), and the female-only analysis of heterogeneous depression attenuated the association (Supplementary Table 10).

PPD also shows a weaker association with SES. This may reflect a more equitable distribution of health services during the postpartum period. Standardised implementation of early detection and management of depressive symptoms in new mothers may reduce the disparity in prevalence seen according to deprivation. While there is some evidence to support this is the case for deprivation, inequalities remain, particularly in ethnic minorities (39).

We found no evidence for a stronger association with BPD PRS in PPD, and do not replicate the findings from Byrne et al. (16) which indicated PPD has a closer genetic relationship to bipolar disorder than depression. The two studies have many differences in methodology, sample characteristics, and phenotype definitions, and our BPD PRS was generated from a larger GWAS, giving more precise estimates of the effect size.

Taken together, these results highlight the importance of considering the likelihood of a depressive episode given the context the participant currently faces. Comparisons across subgroups of depression could provide translational benefit through pointing to risk factors of MD which are directly related with the response to the specified context. Given controls also differ between subgroups, heterogeneity within cases was explored in a case-case analysis which directly tests whether aggregating MD cases can mask associations with a risk factor. As with the subgroup analysis, reported childhood trauma and neuroticism scores were decreased in cases of depression following a chronic disease indicating the prior results are driven partly by heterogeneity within cases. A control-control comparison shows heterogeneity within controls also contributes to our results (Supplementary Table 11). We additionally show an effect of education whereby cases of depression following a chronic disease are more likely to report having no recognised UK based qualifications. Given the association of physical health with SES and that a key separation between the two groups is the requirement for a chronic disease, it is possible that this finding is an SES effect.

We find increased effects for reported childhood trauma in PPD cases relative to heterogeneous depression. Studies have identified PPD as a mediator in the relationship between reported trauma and mother-infant relationships postpartum (40). However, the literature on childhood trauma as a predictor of PPD is sparse and this finding was not significant in the case-control comparison. It is important to stress that trauma was assessed retrospectively, which may differ from prospectively measured trauma (41). An alternative explanation is that becoming a parent leads to greater reflection on both the positive and negative aspects of one’s childhood. This could increase the likelihood of parents finding links between their depressive episode and trauma suffered during childhood relative to non-parents, thus representing mood-congruent recall biases (42).

MD PRS was elevated in PPD cases relative to heterogeneous depression; corroborated by an increased reporting of family history of severe depression. Biological theories often attribute PPD to the same biological changes that occur during pregnancy and the postpartum period. However, a systematic review shows that evidence for these theories are inconsistent (43). While some studies suggest a role for corticotropin-releasing hormone trajectories during pregnancy (44,45), most studies have small sample sizes, are inconsistent in their definition of PPD and do not account for environmental confounders. Our finding suggests biological variation is an important component and that focusing on PPD could improve power in future genetic research for this phenotype. Determining whether PPD has distinct biological pathways will require targeted GWAS such as the ‘Mom Genes’ study (46).

The case-case analysis shows that the weaker association with adulthood trauma and neuroticism observed in the PPD case-control analysis is driven through increased reporting within the control group of PPD (Supplementary Table 11). This also highlights the heterogeneity in control groups, and the importance of clear appropriate definitions. Without subgrouping, meaningful differences may be masked, as shown in the opposite effects between PPD and depression following a chronic disease when using heterogeneous cases act as a reference. Ultimately such differences in risk factors may help to inform stratified treatments.

## Limitations

The findings presented should be considered in light of the following limitations. Firstly, while this study dissects heterogeneity of depression via context, the subgroups themselves remain heterogeneous. For each subgroup, we were unable to determine the severity of the depressive episode which may have contributed to our results. Similarly, the decision to implement a 12-month cut-off for PPD and depression following a chronic disease is arbitrary.

Secondly, for depression following a chronic disease, we aggregated across multiple diseases due to sample size constraints. Our hypothesis was generated through the lens of individual differences in psychological resilience after receiving a diagnosis; resilience may differ by disease and by severity, which we were unable to explore. Other explanations suggest depressive symptoms may arise through shared biology or as side-effects of disease treatment (47).

Thirdly, our phenotypes are derived from self-report questionnaires and electronic health records which each have limitations. Retrospective self-report definitions are subject to recall bias, increasing misclassification of both cases and controls. Electronic health records are prospective but may not represent a complete record of a participant’s medical history. Indeed, HES data relating to psychiatry was not present in Scottish participants meaning potential cases of major depression are missed. Similarly, primary care data is currently only available for approximately 230,000 individuals. Some participants may have experienced a depressive episode without seeking help from a general practitioner. We quantified the impact of data source, showing that effect sizes for most risk factors are comparable between phenotypes of PPD using self-report and electronic health records. Cohort effects from changing prevalence of depression across time may also be present. Portability of PRS across ancestries is poor (48), and our PRS analysis was restricted to those of European ancestries.

Index event bias is a form of collider-stratification bias that occurs when a sample is stratified according to an event which is strongly associated with the risk factor and outcome. Such a bias may therefore manifest in our study design, however, as reasoned in the supplementary discussion, we believe our conclusions are unlikely to be materially impacted.

## Conclusion

We show that the context of a major depressive episode reveals heterogeneity in genetic and epidemiological risk factors. Developments in analytical methods such as natural language processing increasingly afford the opportunity to use the unstructured data present within electronic health records to unpick further contextual subgroups. Integrating such data with structured deep phenotyping present in biobanks may reveal deeper etiological insights through dissecting the heterogeneity of MD.

## Data Availability

Summary statistics used to create bipolar disorder polygenic risk scores (PRS) are made available through the website of the Psychiatric Genomics Consortium at: https://www.med.unc.edu/pgc/download-results/. Major depression PRS contains summary statistics from 23andMe inc. The full GWAS summary statistics for the 23andMe discovery dataset will be made available through 23andMe to qualified researchers under an agreement with 23andMe that protects the privacy of the 23andMe participants. Please visit https://research.23andme.com/collaborate/#dataset-access for more information and to apply to access the data. Individual level data for the UK Biobank is made available to researchers upon application for individual projects.

https://research.23andme.com/collaborate/#dataset-access

## Acknowledgements

CML is funded by the Medical Research Council (MR/N015746). SPH is funded by the Medical Research Council (MR/S0151132).

This study represents independent research funded by the National Institute for Health Research (NIHR) Maudsley Biomedical Research Centre at South London and Maudsley NHS Foundation Trust and King’s College London. The authors acknowledge use of the research computing facility at King’s College London, *Rosalind* (https://rosalind.kcl.ac.uk), which is delivered in partnership with the National Institute for Health Research (NIHR) Biomedical Research Centres at South London & Maudsley and Guy’s & St. Thomas’ NHS Foundation Trusts, and part-funded by capital equipment grants from the Maudsley Charity (award 980) and Guy’s & St. Thomas’ Charity (TR130505). The views expressed are those of the author(s) and not necessarily those of the NHS, the NIHR, King’s College London, or the Department of Health and Social Care.

We thank Dr. Andrew Schork for insightful discussion. We thank participants and scientists involved in making the UK Biobank resource available (http://www.ukbiobank.ac.uk/). UK Biobank data used in this study were obtained under approved application 18177.

