## Supplementary Note for "Context Matters! Depression following childbirth or a chronic disease diagnosis shows specific risk factor profiles"

**Supplementary Information**

**Supplementary Methods**

- **Sub-samples of the UK Biobank for Phenotype Definition**
- **Detecting the first diagnosis of a chronic disease**
- **Definition of incident depression for depression following a chronic disease**
- **Depression According to the CIDI-SF within the Mental Health Questionnaire**
- **Definition of Controls**
- **Exclusion Criteria**

**Supplementary Discussion**

- **The role of trauma in depression following a chronic disease**
- **Explanations for the positive association between level of education and risk for major depression**
- **Index-event bias – what is it and how is it potentially impacting the contextual risk factor associations?**

**Supplementary Figures**

**Supplementary Figure 1: Odds ratio comparison between two definitions of postpartum depression – Electronic Health Records vs Self-report Mental Health Questionnaire.**

**Supplementary Figure 2: Loadings of first two principal components from reported childhood and adulthood trauma.**

**Supplementary Figure 3: Associations between bipolar disorder polygenic risk scores and three contextually based subgroups of major depression.**

**Supplementary Figure 4: Comparison of polygenic risk score odds ratios by p-value threshold.**

**Supplementary Figure 5: Association of bipolar disorder polygenic risk score comparing specific contexts of a depressive episode to a heterogeneous definition of depression.**

**Supplementary Figure 6: Figure 3 of the main text plotted with log odds ratio on the y-axis**

**Supplementary Methods**

**Sub-samples of the UK Biobank for Phenotype Definition**

Information from four complementary datasets were collated to define each depression phenotype. Firstly, the nurse interview, conducted at baseline (2006-2010) by all participants within the UK Biobank (N=502 655) asked about all previous medical diagnoses and treatments (cancer and non-cancer) (1). The same questions were asked for a subset of participants at 3 separate follow-ups in 2012, 2014 and 2019. The Mental Health Questionnaire (MHQ) was completed online by a subset of participants (N=157 366) in 2017. This comprehensive questionnaire focused on retrospective reporting of DSM criteria for numerous psychiatric disorders, as well as a number of well-known questionnaires assessing current symptoms of anxiety and depression (GAD-7, PHQ-9) (2). Hospital records are linked for all participants. These Hospital Episode Statistics (HES) include information on primary and secondary diagnoses for each hospital visit, as well as admission and discharge dates. Dates of linkage are dependent on the country the participant resided in. Record linkage began in England, Wales and Scotland from 1997, 1998 and 1981 respectively. Finally, the primary care data contains record level information about visits to either a general or nurse practitioner**.** This data has been linked for approximately 230 000 participants.

**Detecting the first diagnosis of a chronic disease**

To identify the date of first diagnosis for a selected chronic disease, the earliest recorded hospital admission dates were extracted from primary and secondary diagnoses within the HES dataset. Administrative data is incomplete for each participant, reducing confidence that the earliest record represents the first diagnosis. To overcome this issue, we corroborated the diagnosis using the nurse interview and ensured self-reported age at onset was not prior to the HES data becoming available for a given participant. Cases were retained if participants self-reported the disease, and the self-reported onset was after the date of HES linkage. Self-report diagnoses prior to HES linkage were excluded. If the participant did not corroborate the diagnosis through self-report, they were retained if the earliest HES record was after the date of nurse interview.

**Definition of incident depression for depression following a chronic disease**

Participants were considered not to have had major depression at baseline if they did not report major depression during the nurse interview (field: 20002 – Code 1286), did not report ever having had an antidepressant at baseline (field: 20003, Codes written in Supplementary Table 2) and reported never having been to see a general practitioner or psychiatrist for treatment of nerves, anxiety, tension or depression (fields: 2090 and 2100).

MD diagnoses were collated from primary care records (3), and from HES records relevant to mental health, recorded as either a primary or secondary diagnosis. The earliest date of diagnosis was extracted for each participant (Supplementary Table 3 provides a list of read/ICD codes included).

**Cases of Depression According to the CIDI-SF within the Mental Health Questionnaire**

To be defined as a case of ever having had a major depressive episode, participants had to endorse either of the two cardinal symptoms of major depression - depressed mood and anhedonia (fields: 20446, 20441 respectively). Inclusive of the two cardinal symptoms, five out of a possible eight symptoms had to be endorsed relating to depressed mood, anhedonia, impaired concentration, feeling tired, feelings of worthlessness, change in sleep, weight change in either direction, suicidal thoughts (fields: 20446, 20441, 20435, 20449, 20450, 20532, 20536, 20437 respectively). These symptoms must be accompanied with an impairment to work or social life (field 20440 – responded either ‘somewhat’ or ‘a lot’) and the feelings must have been present for at least ‘most of the day’ (field 20436) for ‘almost every day’ during the episode (field 20439). Age at onset for major depression was identified using field 20433.

**Definition of Controls**

Controls were screened for multiple definitions of depression. Controls did not report depression or antidepressant usage at nurse interview. They were not assigned major depression status (single or recurrent) according to the definition from Smith et al. (4) (field 20126). They had no depression related ICD or read codes from the HES and primary care records respectively. They did not endorse having ever visited a general practitioner or psychiatrist for treatment of nerves, anxiety, tension or depression. For both PPD and depression following a chronic disease, controls must have lived for at least one year following the event.

**Exclusion Criteria**

Cases and controls were excluded if they reported schizophrenia, bipolar disorder or substance abuse (field 20002, codes: 1289 (schizophrenia), 1291 (mania/bipolar disorder/manic depression), 1408 (alcohol dependency), 1409 (opioid dependency), 1410 (other substance abuse dependency)). They reported schizophrenia, any form of psychosis or bipolar disorder in the MHQ (field 20544). They reported usage of an antipsychotic or lithium without an antidepressant (field 20003). They received an ICD code within the HES data relating to substance abuse, schizophrenia or bipolar (F10-F16, F18-F31).

**Genetic Quality Control, Unrelatedness and Ancestry Determination**

Following central quality control (QC) from the UK Biobank (1), ancestry was determined using k-means clustering on the first two principal components of the genetic data with the largest cluster being assumed to correspond to European ancestries.

All single nucleotide polymorphisms (SNPs) had a minimum minor allele frequency of 1% and a call rate > 98%. SNPs were in approximate Hardy-Weinberg equilibrium (p < 10^-8^). Participant self-reported sex was verified using genetic data through X-chromosome homozygosity (FX; FX > 0.9 if male and FX < 0.5 if female) (5). Relatedness was determined using KING (6) whereby a minimum kinship coefficient of 0.44 was imposed. If more than 2 participants were collectively above this threshold - i.e. mother, father and child are all within the UK Biobank sample – an algorithm was employed to retain the largest sample. Finally, genetic principal components were computed on the sample and SNPs remaining after QC using FlashPCA2 (7).

**Risk Factors**

Polygenic Risk Scores (PRS) were computed using the following commands within PRSice v2. Summary statistics from Wray et al. (2018) with UK Biobank samples removed and Stahl et al. (2019) were used to calculate the SNP weights for major depression and bipolar disorder respectively (8,9). Clumping was performed on SNPs within 500kb from the index SNP so that r2 < 0.1 for all SNPs within that window.

In addition to the PRS A set of epidemiological risk factors previously shown to associate with MDD were selected (10): education (reference category = degree; field 6138), socio-economic status (field 189), neuroticism (field 20127), family history of severe depression (11), reported childhood trauma and reported adulthood trauma. Family history of severe depression was taken as the union of the fields 20110 and 20107 which ask about maternal and paternal history of severe depression respectively.

***Education***

Education was adapted from field 6138 which asked ‘Which of the following qualifications do you have?’. Participants could select one or more options from the following options:

- College or University degree
- A levels/AS levels or equivalent (school leaver at age 18)
- O levels/GCSEs or equivalent (school leaver at age 16)
- CSEs or equivalent (school leaver at age 16)
- NVQ or HND or HNC or equivalent
- Other professional qualifications e.g: nursing, teaching
- None of the above
- Prefer not to answer

To reduce the complexity of the question, we first aggregated the responses into a four-level categorical variable corresponding to qualifications attained at (1) college or university (Degree) [reference group], (2) age 18 (A-levels’ or equivalent), (3) age 16 (GCSE or equivalent) and (4) no reported qualifications.

The level ‘age 18 (A-levels’ or equivalent)’ includes all endorsements in the original item for the following categories:

- A levels/AS levels or equivalent (school leaver at age 18)
- NVQ or HND or HNC or equivalent
- Other professional qualifications e.g: nursing, teaching

The level ‘age 16 (GCSE or equivalent)’ includes all endorsements in the original item for the following categories:

- O levels/GCSEs or equivalent (school leaver at age 16)
- CSEs or equivalent (school leaver at age 16)

Prefer not to answer are set to missing.

***Reported childhood trauma***

5 items from the 16-item trauma questionnaire within the MHQ corresponding to trauma occurring during childhood were dichotomised according to the following criteria:

- Field: 20489 - ‘When I was growing up, I felt loved’
  - Endorsed if participant responded ‘Sometimes true’ or worse
- Field: 20491 - ‘When I was growing up, there was someone to take me to the doctor if I needed it’
  - Endorsed if participant responded ‘Sometimes true’ or worse
- Field: 20487 - ‘When I was growing up, I felt that someone in my family hated me’
  - Endorsed if participant responded ‘Sometimes true’ or worse
- Field: 20488 - ‘When I was growing up, people in my family hit me to hard that it left me with bruises or marks’
  - Endorsed if participant responded ‘Rarely true’ or worse
- Field: 20490 - ‘When I was growing up someone molested me (sexually)’
  - Endorsed if participant responded ‘Rarely true’ or worse

***Reported adulthood trauma***

11 items from the 16-item trauma questionnaire within the MHQ corresponding to trauma occurring during adulthood were dichotomised according to the following criteria:

- Field: 20522 - ‘Since I was sixteen, I have been in a confiding relationship’
  - Endorsed if participant responded ‘Sometimes true’ or worse
- Field: 20425 - ‘Since I was sixteen, there was money to pay the rent or mortgage when I needed it’
  - Endorsed if participant responded ‘Sometimes true’ or worse
- Field: 20526 - ‘In your life, have you been in a serious accident that you believed to be life-threatening at the time?’
  - Endorsed if participant responded ‘Any yes’
- Field: 20527 - ‘In your life, have you been involved in combat or exposed to a war-zone (either in the military or as a civilian)‘
  - Endorsed if participant responded ‘Any yes’
- Field: 20528 - ‘In your life, have you been diagnosed with a life-threatening illness?’
  - Endorsed if participant responded ‘Any yes’
- Field: 20529 - ‘In your life, have you been attacked, mugged, robbed, or been the victim of a physically violent crime?’
  - Endorsed if participant responded ‘Any yes’
- Field: 20530 - ‘In your life, have you witnessed a sudden violent death (e.g murder, suicide, aftermath of an accident)?’
  - Endorsed if participant responded ‘Any yes’
- Field: 20521 - ‘Since I was sixteen, a partner or ex-partner repeatedly belitted me to the extent that I felt worthless’
  - Endorsed if participant responded ‘Rarely true’ or worse
- Field: 20523 - ‘Since I was sixteen, a partner or ex-partner deliberately hit me or used violence in any other way’
  - Endorsed if participant responded ‘Rarely true’ or worse
- Field: 20524 - Since I was sixteen, a partner or ex-partner sexually interfered with me, or forced me to have sex against my wishes
  - Endorsed if participant responded ‘Rarely true’ or worse
- Field: 20531 - ‘In your life, have you been a victim of a sexual assault, whether by a stranger or someone you knew?’
  - Endorsed if participant responded ‘Rarely true’ or worse

**Supplementary Discussion**

The purpose of the supplementary discussion is to provide additional insight into the associations between the genetic and epidemiological risk factors and our three contextual definitions of major depression.

**The role of trauma in depression following a chronic disease**

To qualify for a case of depression following a chronic disease, the depressive episode must have been the first experienced by a participant. Further, many of the diseases considered here will arise between mid and late life. An alternative explanation for the decreased association with reported trauma in adulthood and childhood is, therefore, that our definition has removed a subset of cases who experienced their first depressive episode following the onset of an earlier traumatic event. These cases would show a strong association with reported trauma but are excluded from our definition. However, these cases would be included in the heterogeneous depression which may explain the relative increase. Similarly, participants who did not experience a depressive episode following the reported trauma are likely to have increased resilience to future environmental stressors, including the diagnosis of a chronic medical disease. The increased resilience within the contextual subgroup of depression following a chronic disease would likely be reflected as an increased frequency of reported trauma in the controls relative to the general population. Such an explanation highlights the importance of considering environmental stressors across the lifespan as reported trauma associations may shift depending on the type of traumatic event and when the first depressive episode occurs.

**Explanations for the positive association between level of education and risk for major depression**

We observed a positive trend between level of education and risk for MD. Prior literature indicates a complicated relationship between education and MD. Many studies show a protective effect of education (12–14); however, others have highlighted both null (15) and positive associations (16). This variation has been attributed to sample heterogeneity with evidence to support a later age at onset in more highly educated individuals (16) as well as a greater protective effect of education in more disadvantaged participants (14). A known limitation of the UK Biobank is that the participants are not representative of the UK, tending to be of higher SES (17). It is therefore plausible the positive association between MD and level of education is a result of not being able to fully account for participants of low SES. PPD shows differences in the association between each level of education relative to heterogeneous depression, the strongest finding being the protective effect of not having any recognised UK based qualifications relative to having a degree. Sensitivity analyses revealed this association was driven by the definition of PPD taken from the MHQ sample. Given the MHQ is a retrospective assessment across the lifetime, it is possible a greater level of education was correlated with the ability to recall depressive episodes. A higher percentage of total PPD cases were identified using the MHQ, compared to the heterogeneous depression subgroup. It is possible this contributed to the observed differences in the association with level of education (18).

**Index-event bias – what is it and how is it potentially impacting the contextual risk factor associations?**

Index event bias is a form of collider-stratification bias that occurs when a sample is stratified according to an event which is strongly associated with the risk factor and outcome. One of the best used examples of such a bias is known as the smoker’s paradox (19). A study investigated the risk factors associated with mortality following an acute myocardial infarction (AMI). Unexpectedly, smoking was found to be protective of early mortality. The intuitive explanation is that smokers who have suffered from an AMI are likely to be younger and suffer less from other risk factors such as hypertension. This induces a negative association between smoking and the other risk factors for mortality which leads to the observed protective association.

We show via a supplementary analysis that many risk factors explored in this study are associated with giving birth and receiving a diagnosis of a chronic disease (Supplementary Table 12), suggesting the potential for such a bias to manifest. Studies have shown that this bias tends to be relatively low unless the risk factor is very strongly associated with the variable being stratified (20,21). While index-event bias may be contributing partly to the difference in associations, we believe it is not the full explanation given the associations are much lower than those observed for most risk factors and MD. However, we do note the large negative association between level of education and giving birth may play a role in the observed differences to heterogeneous depression.

**Supplementary Figures**

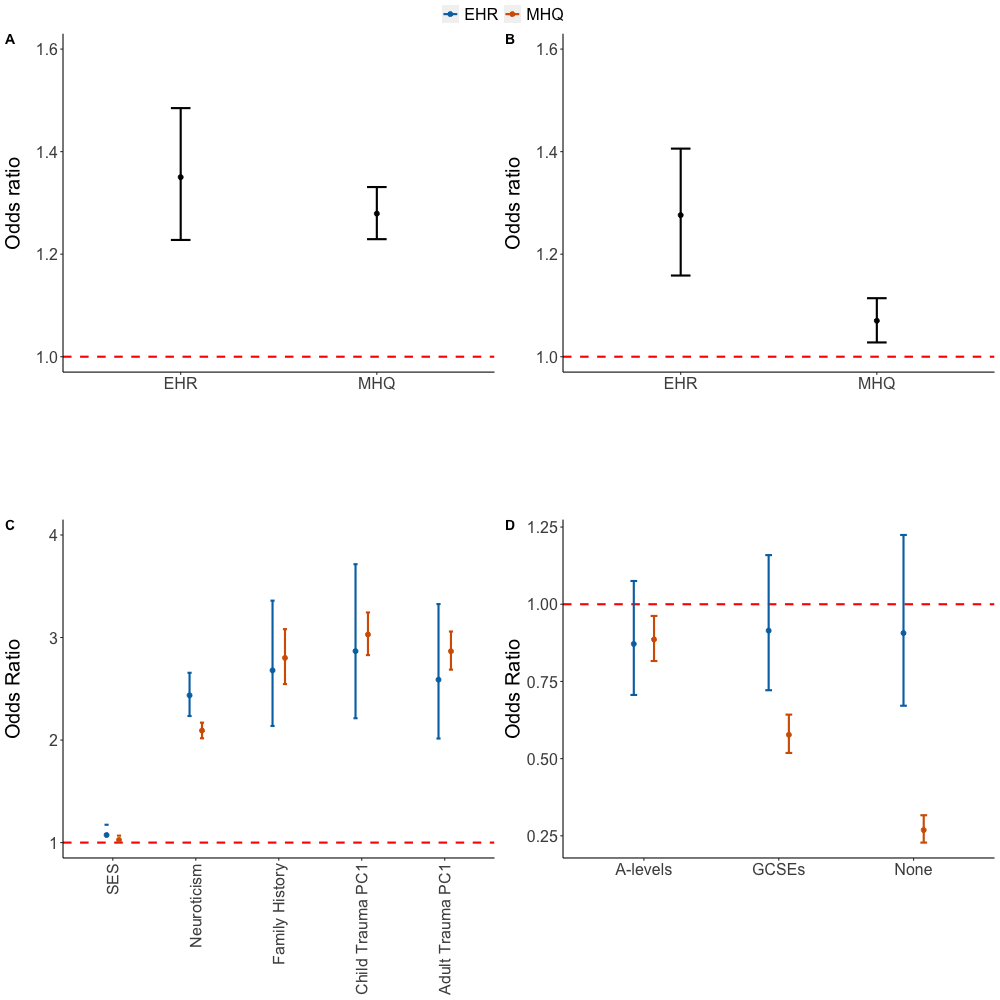

**Supplementary Figure 1: Odds ratio comparison between two definitions of postpartum depression – Electronic Health Records vs Self-report Mental Health Questionnaire.** A.) MDD PRS odds ratio. B.) BPD PRS odds ratio. C.) Epidemiological risk factors odds ratio. D.) Level of education odds ratio. All error bars represent 95% confidence intervals.

**A.**

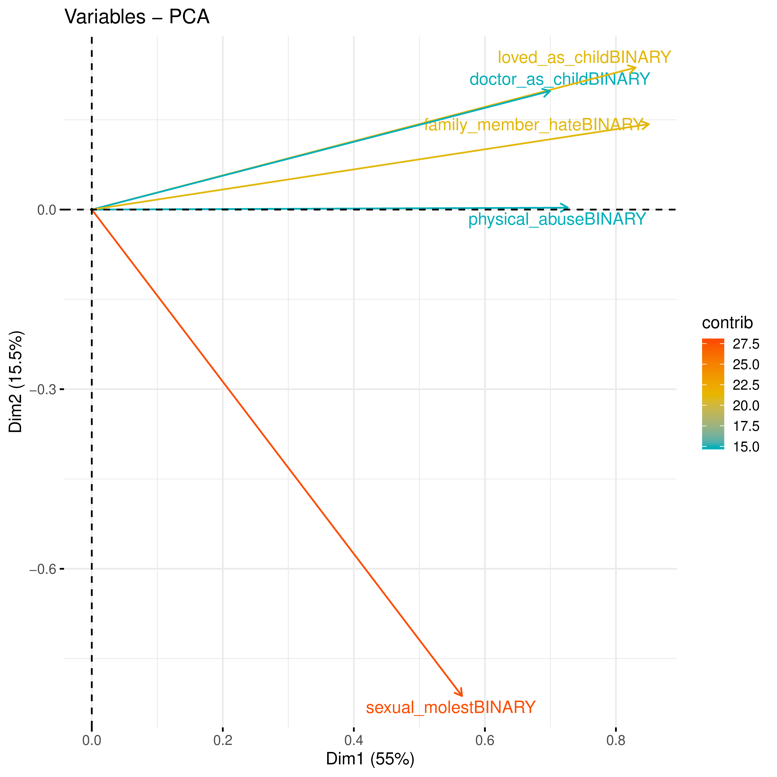

**B.**

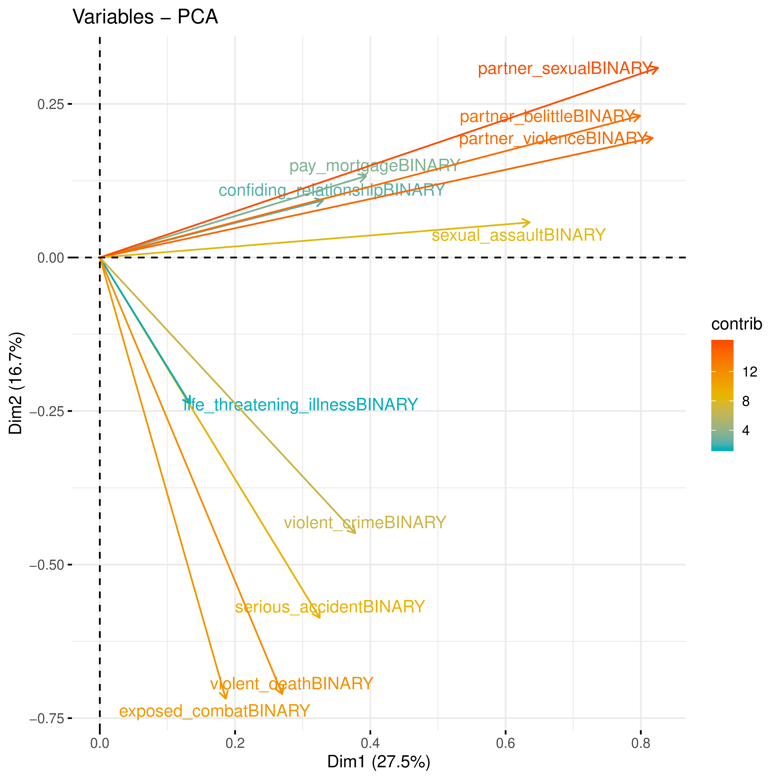

**Supplementary Figure 2: Loadings of first two principal components from reported childhood and adulthood trauma.** A.) Reported childhood trauma items. B.) Reported adulthood trauma items.

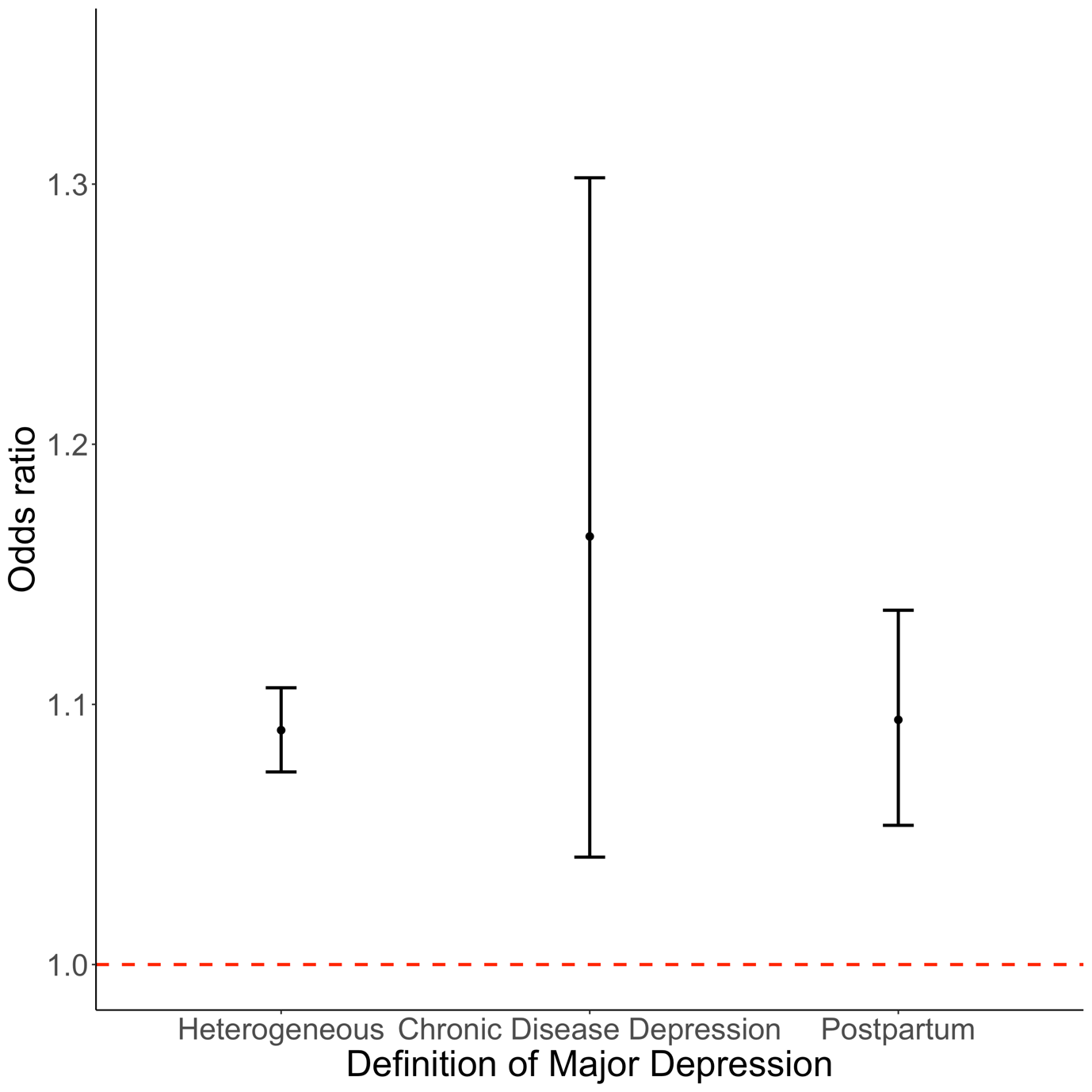

**Supplementary Figure 3: Associations between bipolar disorder polygenic risk scores and three contextually based subgroups of major depression.** Error bars represent 95% confidence intervals.

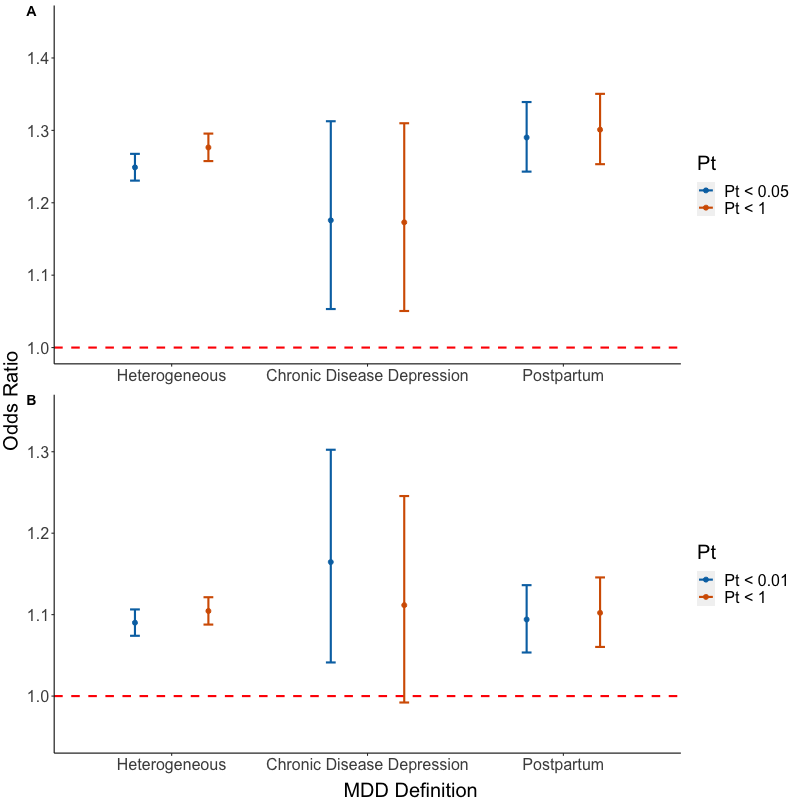

**Supplementary Figure 4: Comparison of polygenic risk score odds ratios by p-value threshold.** In this figure, the original thresholds, Pt < 0.05 and Pt < 0.01, for major depression and bipolar disorder respectively are compared to a threshold which includes all single nucleotide polymorphisms (SNPs) to verify that this choice did not materially change results. A.) Comparison for major depression polygenic risk scores. B.) Comparison for bipolar disorder polygenic risk scores. Error bars represent 95% confidence intervals.

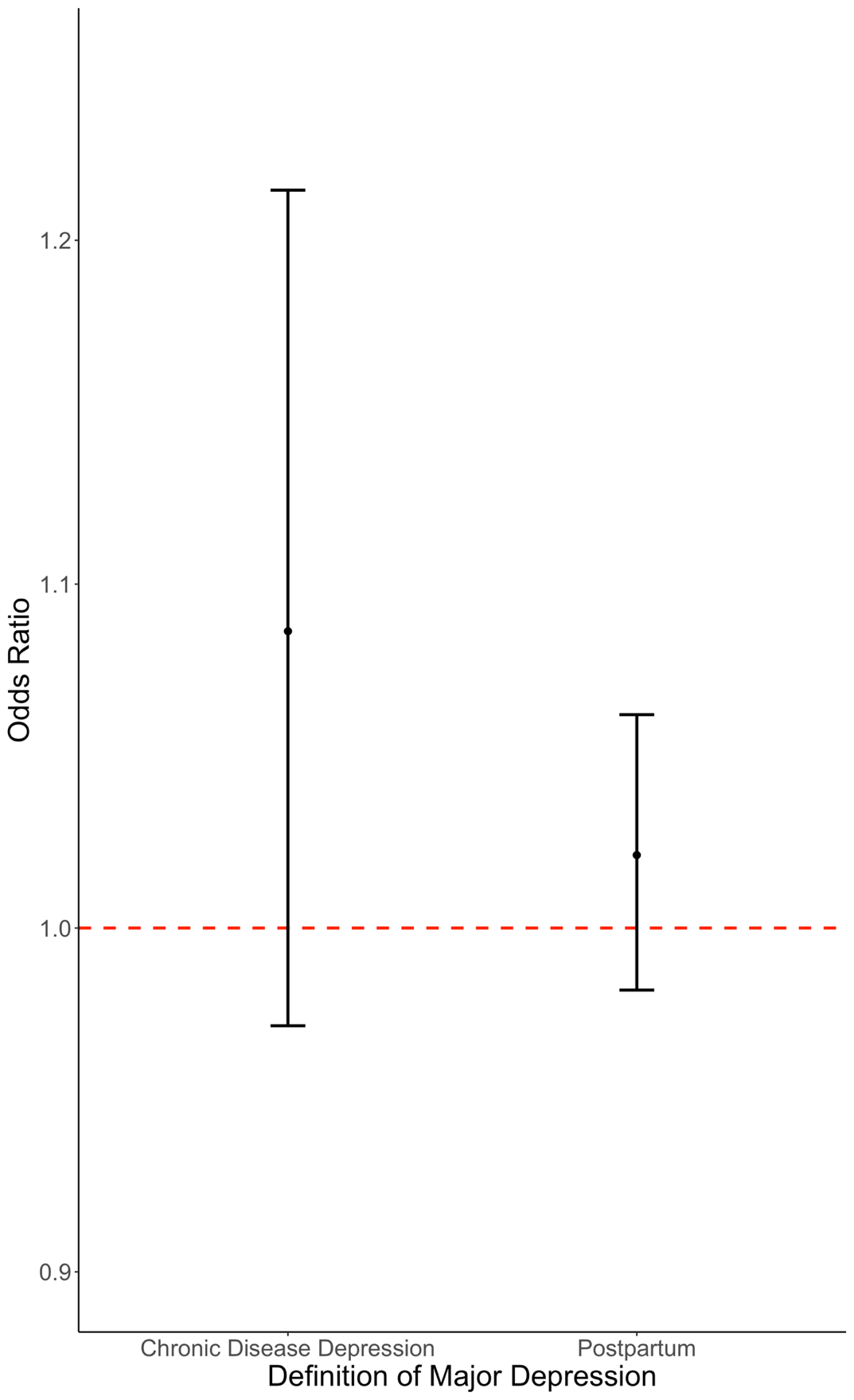

**Supplementary Figure 5: Association of bipolar disorder polygenic risk score comparing specific contexts of a depressive episode to a heterogeneous definition of depression.** In each test, cases within the heterogeneous depression subgroup are the reference group. Error bars represent 95% confidence intervals.

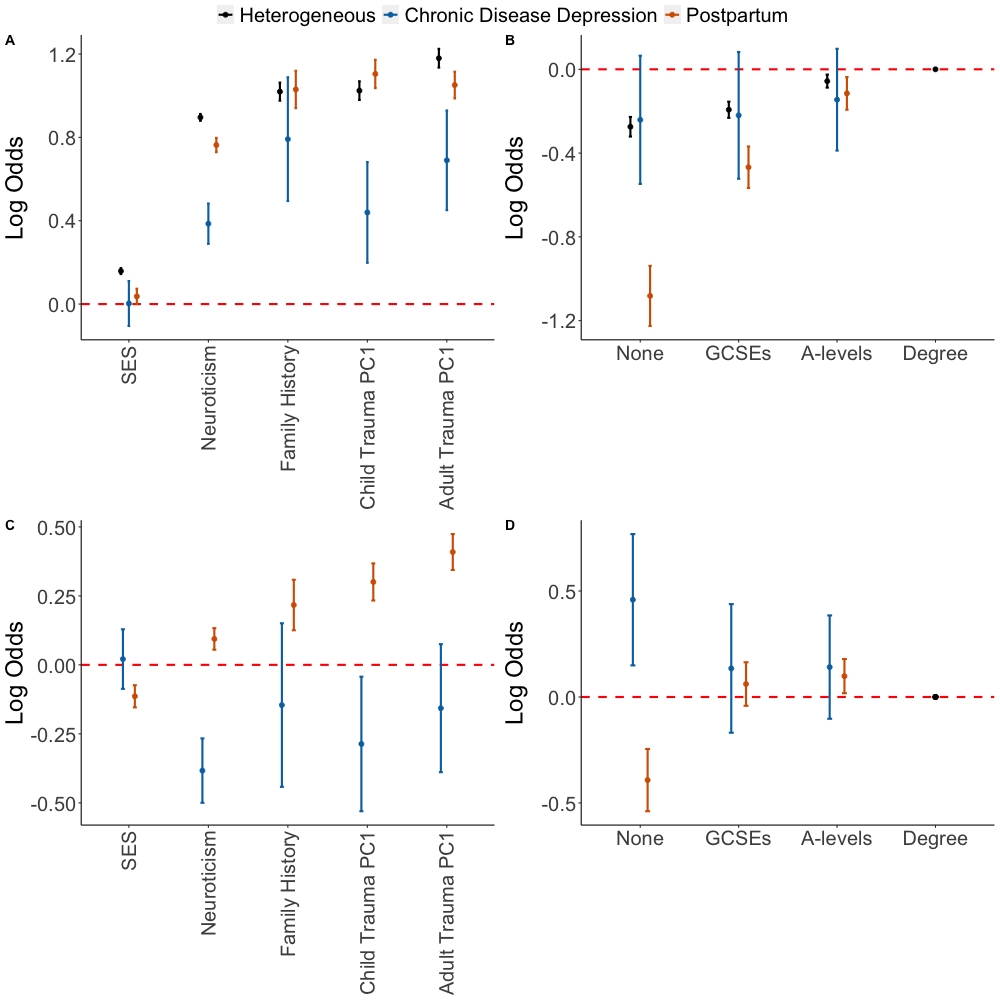

**Supplementary Figure 6: Figure 3 of the main text plotted with log odds ratio on the y-axis. This is to show the differences between the risk factors on a linear scale. A) Association of three contextually based subgroups of major depression with epidemiological risk factors. B) Association of three contextually based subgroups of major depression with level of education.** Having a college or university degree is the reference category**. C) Case-case comparisons for epidemiological risk factors. D) Case-case comparison for educational attainment with all individuals with a university degree as the reference.** In each case-case comparison, cases of heterogeneous depression are the reference group. For all graphs error bars represent 95% confidence intervals. The dashed red line represents the point at which the risk factor shows no association with the contextual definition of major depression (log odds = 0).
